# Toward optimized intravoxel incoherent motion (IVIM) and compartmental T2 mapping in abdominal organs

**DOI:** 10.1101/2025.07.14.25331475

**Authors:** Julia Stabinska, Thomas A. Thiel, Hans-Jörg Wittsack, Alexandra Ljimani, Helge J. Zöllner

## Abstract

**Purpose:** To quantitatively assess the bias in the intravoxel incoherent motions (IVIM)-derived pseudo-diffusion volume fraction (*f*) caused by the differences in relaxation times between the tissue and fluid compartments, and to develop a 2D fitting approach and an optimal acquisition protocol for the relaxation compensated T2-IVIM imaging in the liver and kidney.

**Methods:** Numerical simulations were conducted to investigate the TR- and TE-dependent bias in *f* when using the conventional IVIM model, and to evaluate the applicability of the extended 2D T2-IVIM model for reducing this bias. The *in silico* findings were then validated using the *in vivo* IVIM data from healthy volunteers on a clinical 3-Tesla MRI scanner. Finally, a numerical framework for optimizing the T2-IVIM protocol for relaxation-compensated *f* parameter estimation was proposed and tested using *in vivo* data.

**Results:** When using the traditional IVIM model, a trend toward higher *f* with increasing TE was found in the liver (R = 0.42, P = 0.043), but not in the kidney cortex (R = −0.067, P = 0.76) and medulla (R = 0.039, P = 0.86). The 2D T2-IVIM modeling yielded lower *f* and reduced the intra-subject variability in the liver. Our results also suggest that a b-TE protocol with six b-values and three different TE values (50, 55, and 100 ms) might be optimal for liver T2-IVIM.

**Conclusion:** The extended 2D T2-IVIM model combined effectively minimizes the TE-dependent bias in *f* and allows simultaneous estimation of the IVIM parameter and compartmental T2 values in the liver and kidney.

## Introduction

Molecular diffusion in biological tissues is influenced by both microstructure and microdynamics, including capillary blood flow, exchange, and transport between different tissue compartments^1^. Diffusion-weighted MRI can, therefore, provide information on the hemodynamic status of tissues and detect changes in microvascular perfusion related to both physiological (e.g., response to stimuli or exercise^2–4^) and pathological conditions (e.g., stroke^5,6^, cancer^7–11^, and kidney^12–16^ and liver^17–20^ diseases).

As postulated by Le Bihan et al. in 1980s^21,22^, blood flow within isotropically distributed capillary segments induces phase dispersion of the MR signal, which results in enhanced signal attenuation in diffusion-weighted images, especially at lower b-values. To separate the contributions of “true” molecular diffusion and microcirculation of blood in the capillaries, they proposed a method called “intravoxel incoherent motion (IVIM) imaging” that relies on acquiring multi-b-value DWI data. The IVIM model assumes that (i) the pseudo-diffusion coefficient associated with microcirculation is substantially higher than the molecular diffusion coefficient, (ii) exchange between vascular and extracellular compartments is negligible during measurements, and (iii) relaxation times of the involved compartments are identical. Under these conditions, the signal decay across different b-values is expected to follow a biexponential function:

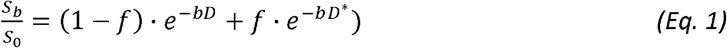

Where *f* is the pseudo-diffusion volume fraction (commonly referred to as the perfusion fraction), *D* and *D\** are the “true”- and pseudo-diffusion coefficients, and S_b_ and S_0_ are the signal intensities measured with gradient sensitivity factor *b* and *b* = 0, respectively.

The IVIM data is typically acquired using a single echo time, usually the minimum, dictated by the duration needed to include the largest diffusion gradient alongside the spin-refocusing pulse and imaging redout. The TE and TR dependencies are then generally incorporated into the S_0_ term during the data fitting step. However, since both IVIM model compartments exhibit distinct and variable T1 and T2 relaxation times, the traditional IVIM model is prone to inaccuracies in estimating the pseudo-diffusion volume fraction^22^. In particular, as body fluids such as blood and pre-urine are known to have longer T2 than most tissues^23^, *f* tends to be overestimated as a function of increasing TE^24^, by a factor proportional to the difference in T2 between the tissue and fluid components^22^. If unaccounted for, this TE-dependence of *f* has negative implications, especially for clinical trials in which the IVIM imaging is performed and/or repeated on different scanners.

In pursuit of standardizing the use of IVIM in clinical studies, in this study we aim (i) to quantitatively assess the effect of relaxation time differences between tissue and flow-related compartments on the IVIM parameter estimation, (ii) to develop a two-dimensional IVIM data fitting approach for simultaneous estimation of the IVIM parameters and compartmental T2 values, and (iii) to provide a generalized numerical framework for optimizing the b-TE protocol for IVIM imaging. Our findings are validated using *in vivo* IVIM data collected in the kidneys and liver of healthy volunteers on a clinical 3T MRI system.

## Methods

*In silico* DW signal generation, data fitting, analysis and visualization were performed using in-house developed scripts in MATLAB (2024b, Mathworks, MA) and Python 3.10 and are freely available on Github: https://github.com/stabinska/T2-IVIM.

### Numerical phantom

Diffusion-weighted MR signals were generated using the modified relaxation-compensated IVIM equation:

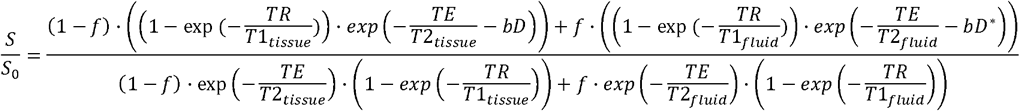

where T1_tissue_, T2_tissue_ and T1_fluid_, T2_fluid_ are the longitudinal and transverse relaxation times for the tissue and fluid (e.g., blood and/or pre-urine in the kidney) compartments, respectively. The DW signals were generated using biologically realistic estimates of IVIM parameter values and different ranges of T1 and T2 relaxation times at 3T. Typical IVIM acquisition parameters with 6 typical b-values: (0, 10, 100, 200, 500, and 800 s^2^/mm) and broad ranges of TE (47 – 92 ms in 5 ms steps) and TR values (2000 – 4000 ms in 500 ms steps) were chosen (“General” phantom in Table 1). Monte-Carlo simulations were performed by generating 2500 repetitions of each parameter combination with five different levels of Rician noise SD: 0 and 0.025. A summary of all simulation parameters is given in Table 1.

**Table 1:** Tissue and acquisition parameters used for numerical simulations.

| Parameter | Value |  |
| --- | --- | --- |
| **Phantom type** | General | Liver at 3T |
| **f (%)** | 23% | 9.5% |
| **D (mm<sup>2</sup>/s)** | 0.0011 | 0.001 |
| **D\* (mm<sup>2</sup>/s)** | 0.07 | 0.067 |
| **T1<sub>tissue</sub> (ms)** | 800 | 800 |
| **T1<sub>fluid</sub> (ms)** | 800 to 1400 in 50 ms steps | 800 |
| **T2<sub>tissue</sub> (ms)** | 27 | 27 |
| **T2<sub>fluid</sub> (ms)** | 27 to 87 in 5 ms steps | 27 to 87 in 5 ms steps |
| **b-values (s/mm<sup>2</sup>)** | 0,10, 100, 200, 500, and 800 | 0, 10, 20, 100, 200, and 550 |
| **TE (ms)** | 47 to 92 in 5 ms steps | TE set 1: 47 to 72 in 5 ms steps<br>TE set 2: 50 to 100 in 5 ms steps |
| **TR (ms)** | 2000, 2500, 3000, 3500, 4000 | 4000 |
| **SD<sub>Rician noise</sub>/SNR at S<sub>0</sub> (SNR<sub>S0</sub>)** | 0/∞, 0.025/40 | 0.025/40 |
| **Repetitions (N<sub>rep</sub>)** | 2500 | 1 |

### MRI data fitting

The generated DW signals were subsequently fitted using the conventional IVIM model (Eq. 1) for each TE separately to determine the IVIM parameters (S_0_, D, D* and f) (*1D IVIM fit*), and the extended two-dimensional T2-IVIM modelling (*2D T2-IVIM fit*) simultaneously including all six TE values and all six b-values to estimate the IVIM parameters as well as the T2_tissue_ and T2_fluid_ values (**Figure 1**) using a T2-relaxation compensated IVIM equation. The IVIM parameters and compartmental T2 values were determined simultaneously by reparametrizing the two-dimensional data and modelling as a one-dimensional problem. The bias (in %) in the IVIM parameter estimates was calculated as 100·(*f*_*fit*_ – *f*_*true*_)/f_true_.

**Figure 1.**
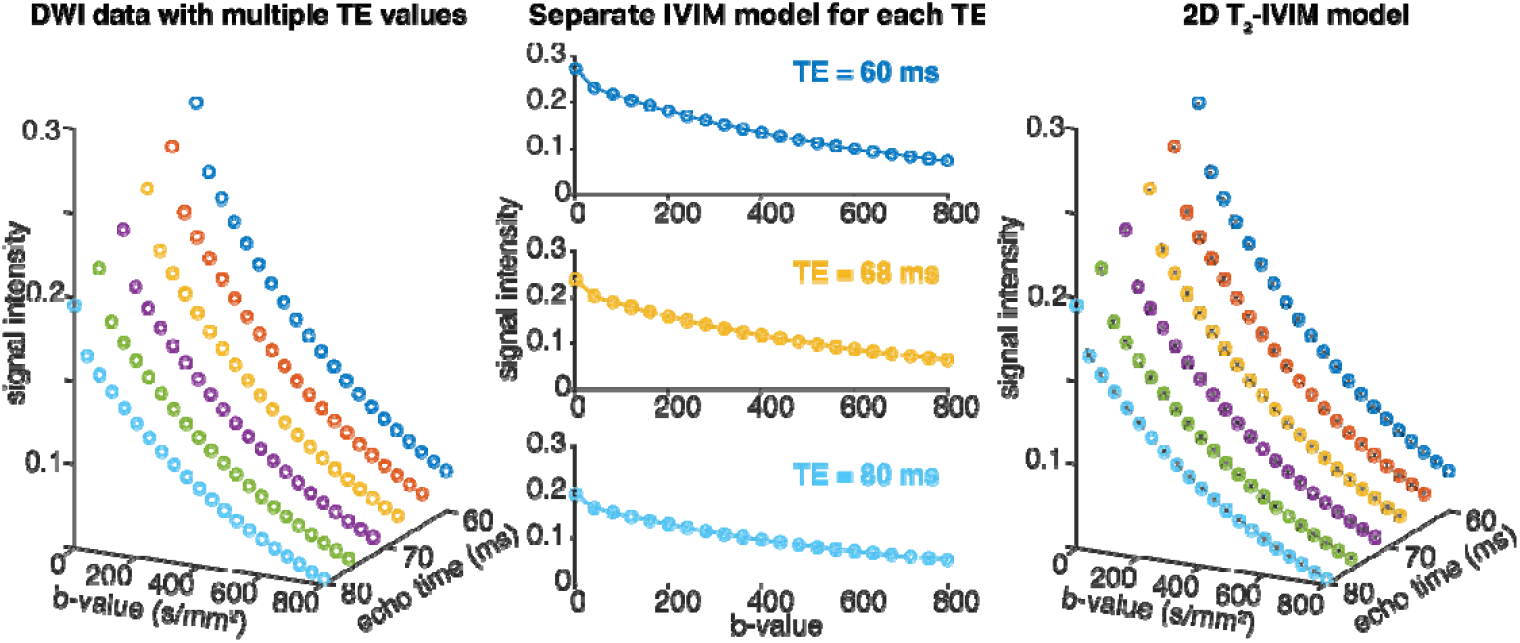
A schematic diagram of data modelling approaches used in this study. DW signals at different TE (and TR) and b-values were generated (left panels). These signals were then fitted using the conventional IVIM model for each TE separately (middle panel), and the extended 2D T2-IVIM modelling for all combinations of TE and b-values. DW signals at five different TE values and a single TR = 3000 ms are depicted for visualization purposes.

### In vivo MRI experiments

The study was approved by the local ethics committee, and written consent was obtained from each participant before the MRI examination. Four healthy subjects (3 females and 1 male; age range: 25 - 30⍰years; mean age: 26.5 ± 3.1⍰years) with no history of renal or liver diseases were included in this study. Subjects were not given any restrictions regarding fluid or food intake.

MRI examinations were conducted on a 3T MRI system (MAGNETOM Prisma; Siemens Heathineers, Erlangen, Germany) using an 18-channel torso array coil and a 32-spine coils. For anatomical visualization and planning, T2 HASTE (half Fourier single-shot turbo spin-echo) coronal oblique images were acquired. DWI data were collected using a DW monopolar pulsed gradient spin-echo (PGSE) sequence with the following parameters: TR = 1900⍰ms; FOV: 370⍰×⍰370⍰mm^2^; acquisition matrix: 176⍰×⍰176; voxel size: 2.1⍰×⍰2.1⍰×⍰5.0⍰mm^3^; bandwidth: 1894⍰Hz/Px, GRAPPA factor: 2; Phase Partial Fourier: 5/8; 3 slices; 3 diffusion directions (3D diagonal); b-values (averages): (0 (3), 10 (3), 20 (3), 30 (3), 50 (3), 70 (4), 100 (4), 150 (4), 200 (4), 250 (4), 300 (4), 350 (4), 450 (5), 550 (5), 650 (5), 750 (5)) s/mm^2^, and spectral-attenuated inversion-recovery fat saturation for fat suppression. This sequence was repeated at six different TE values = (47, 52, 57, 62, 67, and 72) ms. The diffusion time (Δ = 20 ms) and diffusion gradients duration (*δ* = 6.2 ms) were kept constant at each TE to minimize their influence on the IVIM parameter^25^. The DWI acquisition was respiratory-triggered at the exhale phase of the respiratory cycle with a 20% threshold. Depending on the subject’s respiration rate, the average acquisition time for each DWI acquisition was about 4 – 6 minutes.

### In vivo data analysis

Retrospective two-dimensional registration based on a normalized mutual information similarity measure was performed to align all DW images acquired at different TE- and b-values using an in-house-developed software based on ANTs (http://stnava.github.io/ANTs). After the motion correction, a 3 × 3 Gaussian filter was applied to each image. Following a visual inspection by an experienced abdominal radiologist (A.L., 10 years of experience), a single slice that was least affected by respiratory motion and contained clearly visible kidney cortex and medulla in S_0_ image was selected for manual ROI segmentation (kidney cortex, kidney medulla, and liver) in ITK-SNAP^26^ (version 3.8.0) and further analysis.

The *in vivo* data was fitted pixel-wise using the traditional IVIM model for each TE separately and using the 2D T2-IVIM model. To improve the robustness of compartmental T2 and IVIM parameter estimation, modified versions of the conventional segmented (“two-step”)^27,28^ and the Image Downsampling Expedited Adaptive Least-Squares (IDEAL)^29,30^ approaches were applied. With the two-step method, a single T2 value was fitted in the first step and subsequently used as initial guess for the T2_slow_ parameter estimation in the second step. For the IDEAL approach, 11 downsampling steps were applied (1 × 1, 2 × 2, 4 × 4, 8 × 8, 16 × 16, 32 × 32, 64 × 64, 96 × 96, 128 × 128, 152 × 152, and 176 × 176) and the initial guesses and boundaries for each fitting step were constrained to 50% of the initial values for S_0_, and 20% for f, D and D*. The initial parameter values in each iterative step were determined by spatially interpolating the estimated parameters from the prior downsampled image using bilinear interpolation.

### Numerical TE-b protocol optimization and in vivo validation

Due to the tissue-specific differences in diffusion and relaxation parameters, the TE-b protocol optimization for T2-IVIM imaging was performed separately for the kidney and liver. All parameters used for the numerical kidney and liver phantoms can be found in Table 1.

The optimal sets of TE values for T2-IVIM were found using a genetic algorithm solver (*ga* function in the Global Optimization Toolbox, MATLAB), which is a stochastic, population-based algorithm for solving both constrained and unconstrained optimization problems that is based on a natural selection process. In each generation, the genetic algorithm selects a set of individuals from the current population to be “parents” and uses them to produce “children” for the next generation. Over successive generations, the population “evolves” toward an optimal solution through mutations and crossover among population members^31^. The genetic algorithm offers several advantages over the least-squares solvers, particularly in complex and non-linear optimization problems, including higher robustness against finding local minima in large search spaces^32^. Two TE sets were considered, one containing the TE values used for the *in vivo* experiments (TE set 1: 47 – 72 ms in 5 ms steps) and one with a broader range of TE values (TE set 2: 50 – 100 ms in 5 ms steps). The genetic algorithm was used to minimize the normalized root-mean-square-error (nRMSE) of the pseudo-diffusion volume *f*:

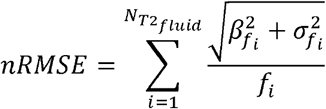

with the bias β and standard deviation σ for a given pseudo-diffusion volume fraction, *f*_*i*_ summed across the different T2_fluid_ values. The normalization ensures that relative deviations not absolute are considered. This way the coefficient of variation is minimized instead of the absolute standard deviation of *f*. The variance was estimated using the Cramer-Rao Lower Bound (CRLB) of the pseudo-diffusion fraction f to improve the speed of the optimization process. The optimization was repeated to find the optimal TE values for a 2, 3, 4, and 5 TEs TE-b protocol for the TE set 1 and TE set 2. The b-values were adopted directly from the consensus guidelines for IVIM without further optimization in this study. TR was kept constant at 4000 ms. For the genetic algorithm the population size was set to 10,000 with 100 total generations and a function tolerance of 0.001. The TE values from the fittest 5% of the population were preserved between generations.

Results of the protocol optimization for the TE set 1 were tested for the liver using a subset of the in vivo data at b-values = (0, 10, 100, 200, 550, and 750) s/mm^2^ and different sets of the optimal TE values. To evaluate the relative bias in *f*, a difference Δ*f* map was calculated by subtracting the *f* map obtained with each TE-b protocol from the ‘ground truth’ *f* map obtained using the full TE-b protocol (6 TEs and 16 b-values).

### Statistical analysis

The ROI-based IVIM parameter estimates obtained from the 2D T2-IVIM and conventional IVIM modeling for each TE value were compared using planned pairwise ANOVA comparisons with Bonferroni correction with an adjusted significance level of *α* = 0.00833 (= 0.05/6). The relationship between the IVIM parameters and TE from the conventional IVIM modeling was assessed using the Spearman correlation with a significance level *α* = 0.05.

## Results

### Effects of relaxation time differences between the IVIM compartments on parameter estimation

As shown in **Figure 2**, pseudo-diffusion volume fraction *f* strongly depends on the differences in relaxation times (ΔT1 = T1_fluid_ – T1_tissue_ and ΔT2 = T2_fluid_ – T2_tissue_) between both compartments, and thus on selected TE and TR. In fact, even at a relatively short TE of 52 ms, the bias in *f exceeds 20% for* Δ*T2 as small as 5 ms. The effect of differences in compartmental T1 on the accuracy of f parameter estimation is less pronounced and can be minimized by choosing a longer TR (>3000 ms)*.

**Figure 2.**
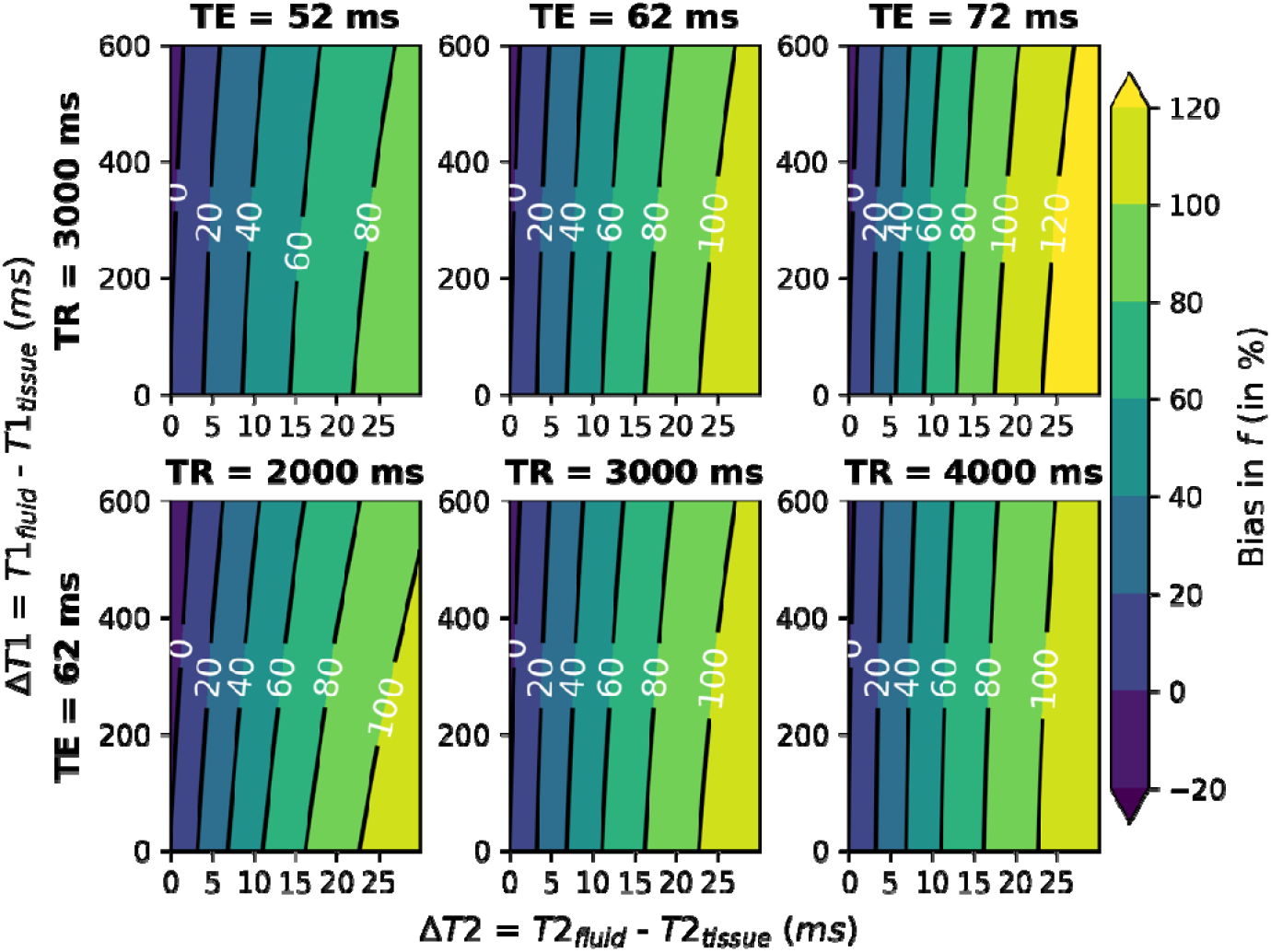
Bias in pseudo-diffusion volume fraction *f* (%) as a function of differences in compartmental relaxation times at different TE and TR values generated without Rician noise.

*As demonstrated in Figure 3, an extended 2D T2-IVIM model that allow for distinct T2 values in both compartments can be used to efficiently minimize the bias in f estimates for* Δ*T2 > 30 ms and over a wide range of* Δ*T1 values (>600 ms)*.

**Figure 3.**
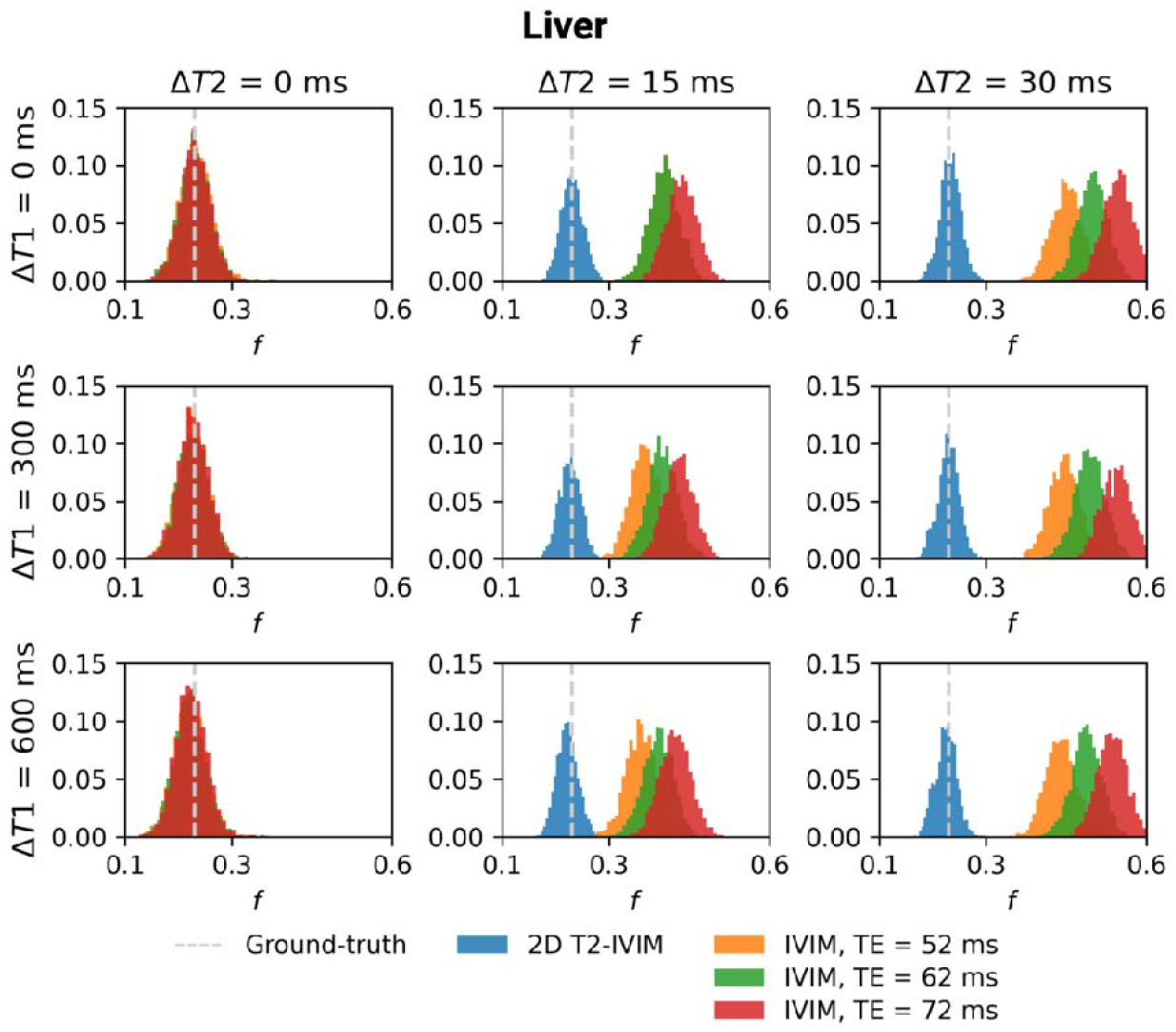
Distributions of pseudo-diffusion volume fraction *f* obtained from the 2D T2-IVIM modeling (blue), and conventional IVIM modeling at TE = 52 ms (yellow), TE = 62 ms (purple), and TE = 72 ms (red) using Monte-Carlo simulations with N_rep_ = 1000 repetitions and SNR_S0_ = 40. TR = 4000 ms was kept constant.

Figure 4. shows exemplary T2 and IVIM parameter maps in the kidney and liver of a healthy volunteer obtained from the data fit to the conventional IVIM model (at TE = 47 ms) and 2D T2-IVIM model with the segmented (top row) and IDEAL approaches (middle and bottom rows). The latter fitting method produced more visually appealing maps than the former one, which agrees with our prior work^29^. The pseudo-diffusion fraction maps show clear differences between the IVIM and T2-IVIM models with overall higher *f* values in regions with high flow fractions, like the renal pelvis and the liver. As expected, the differences in diffusion coefficient maps between the 1D and 2D models are negligible. At the same time, slightly decreased pseudo-diffusion D* values can be observed in the relaxation-time compensated maps, which can partially be explained by the overall higher variability in estimating this parameter.

The results of the ROI-based group analysis are demonstrated in **Figure 5** and in the Supplementary Material **Figure S1**. A trend towards higher *f* with longer TE can be observed in the liver for the traditional IVIM model (R = 0.42, P = 0.043), which is consistent with our numerical findings. While generally not statistically significant (P > 0.0083 for all comparisons, except for f_T2-IVIM_ vs f_IVIM_ at 67 ms, where P = 0.0076), this effect appears to be largely reduced when 2D T2-IVIM modeling was applied, leading to overall decreased *f* values and lower variability between the subjects compared to the IVIM modeling. Interestingly, no such trend was observed in the kidney in the considered TE values range (P > 0.17 for all comparisons).

**Figure 4.**
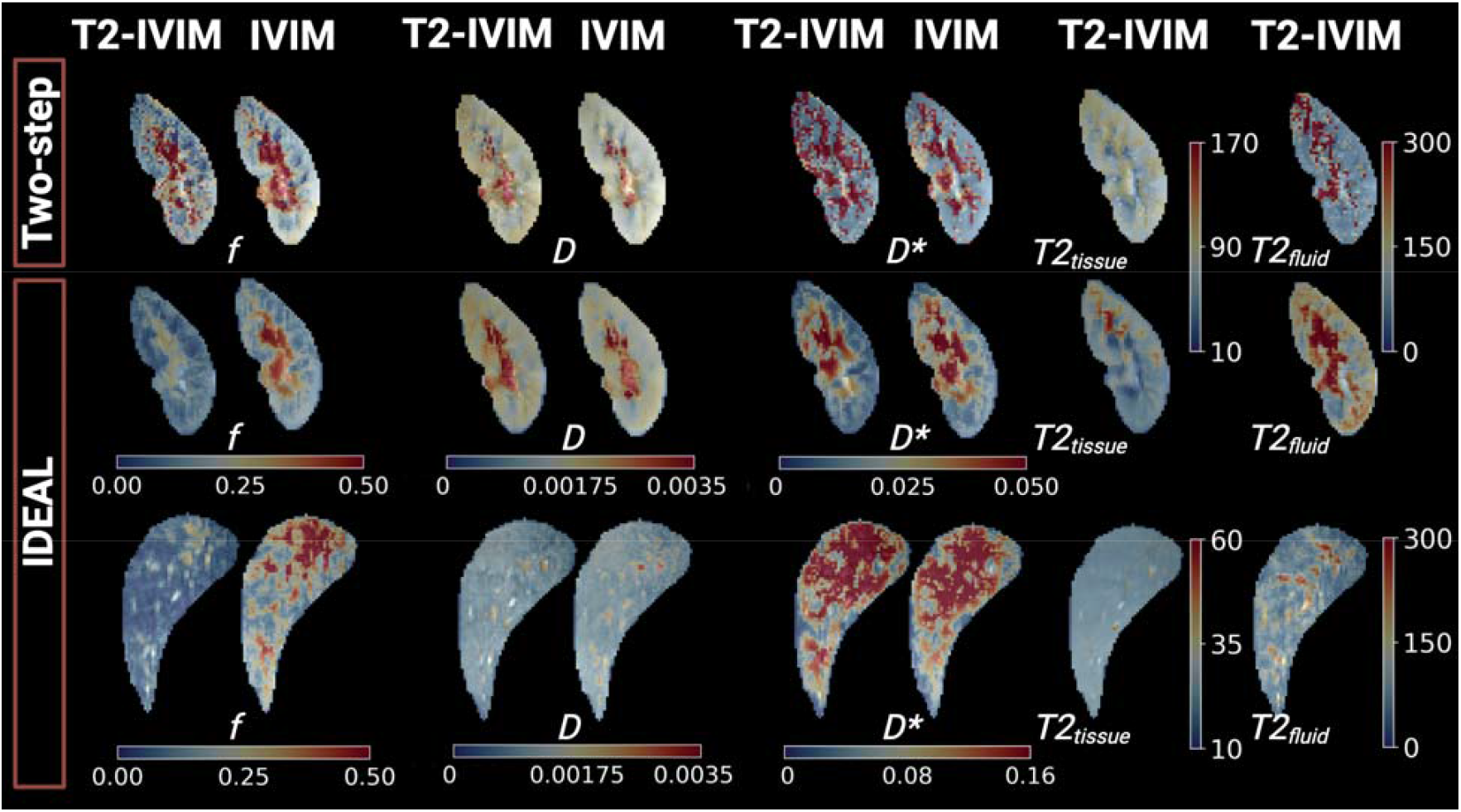
Representative IVIM parameter maps obtained in the kidney and liver of a healthy volunteer using the conventional IVIM (at TE = 47 ms) and 2D T2-IVIM modelling.

**Figure 5.**
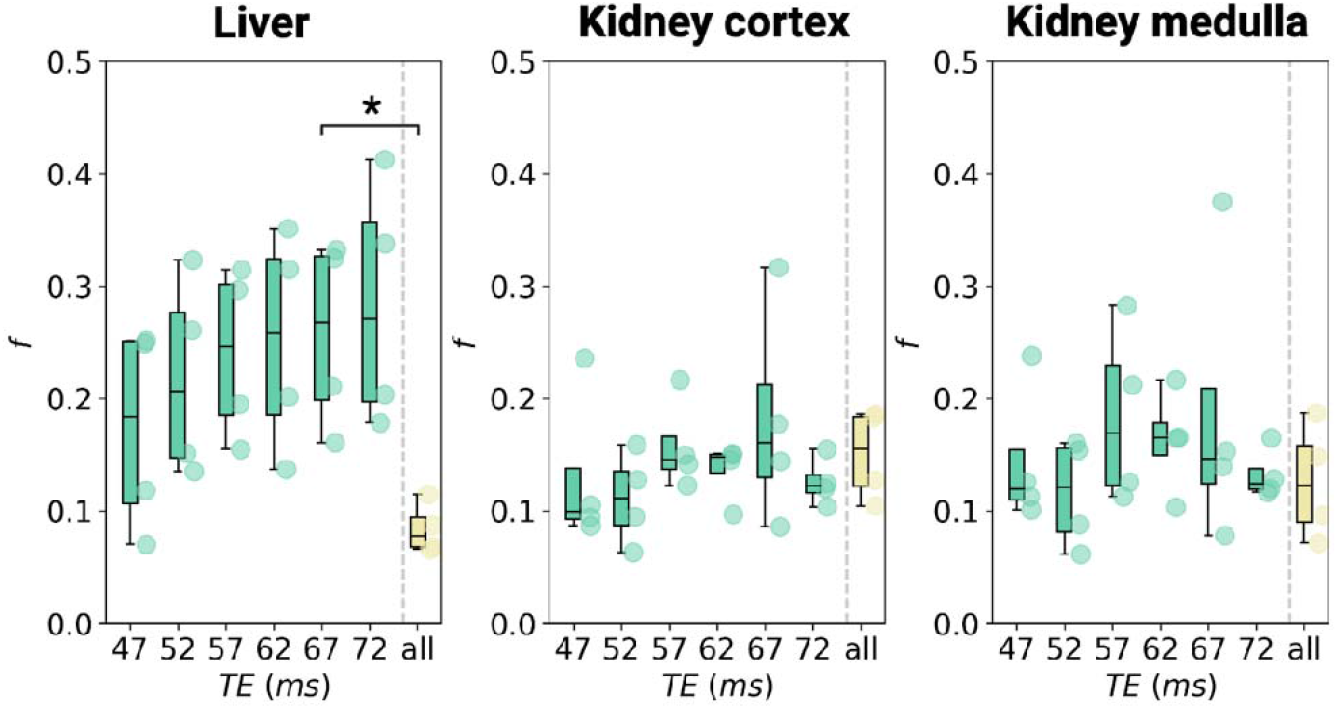
Pseudo-diffusion volume fraction *f* values obtained in all volunteers (N = 4) in the liver, kidney cortex, and kidney medulla using the IVIM modeling for each TE separately (green) and simultaneous 2D T2-IVIM modeling (yellow). The results for each volunteer are displayed as a circle. (*) P < 0.0083.

Furthermore, no significant correlations between f and TE were found neither in the kidney cortex (R = −0.067, P = 0.76) nor kidney medulla (R = 0.039, P = 0.86). Similarly, there was no difference in D and D* obtained from the 2D T2-IVIM and IVIM fitting (P > 0.05 for all comparisons) and no significant correlations between these parameters and TE (all P > 0.05) in neither organ (**Figure S1**).

The results of the numerical optimization for the TE set 1 and set 2 in the liver are displayed in **Figure 6A and 6B**, respectively. For both TE sets, the minimal and maximal TEs are the common optimum regardless of the number of TEs in the protocol. For the TE set 1, the distributions of *f* values obtained using the T2-IVIM model with the optimal TE values show that increasing the number of TEs improves the fitting performance; however, the difference between the two-TEs and three-TEs protocols is comparatively negligible. After extending the TE range in the TE set 2, a shift toward higher optimal TE values for all sets can be observed. The distributions of *f* values become sharper, and the accuracy of the estimation of *f* is also improved. In general, a protocol with two shorter (50 and 55 ms) and one longer TE (100 ms) appears to be a suitable compromise between accuracy and scan time.

**Figure 6.**
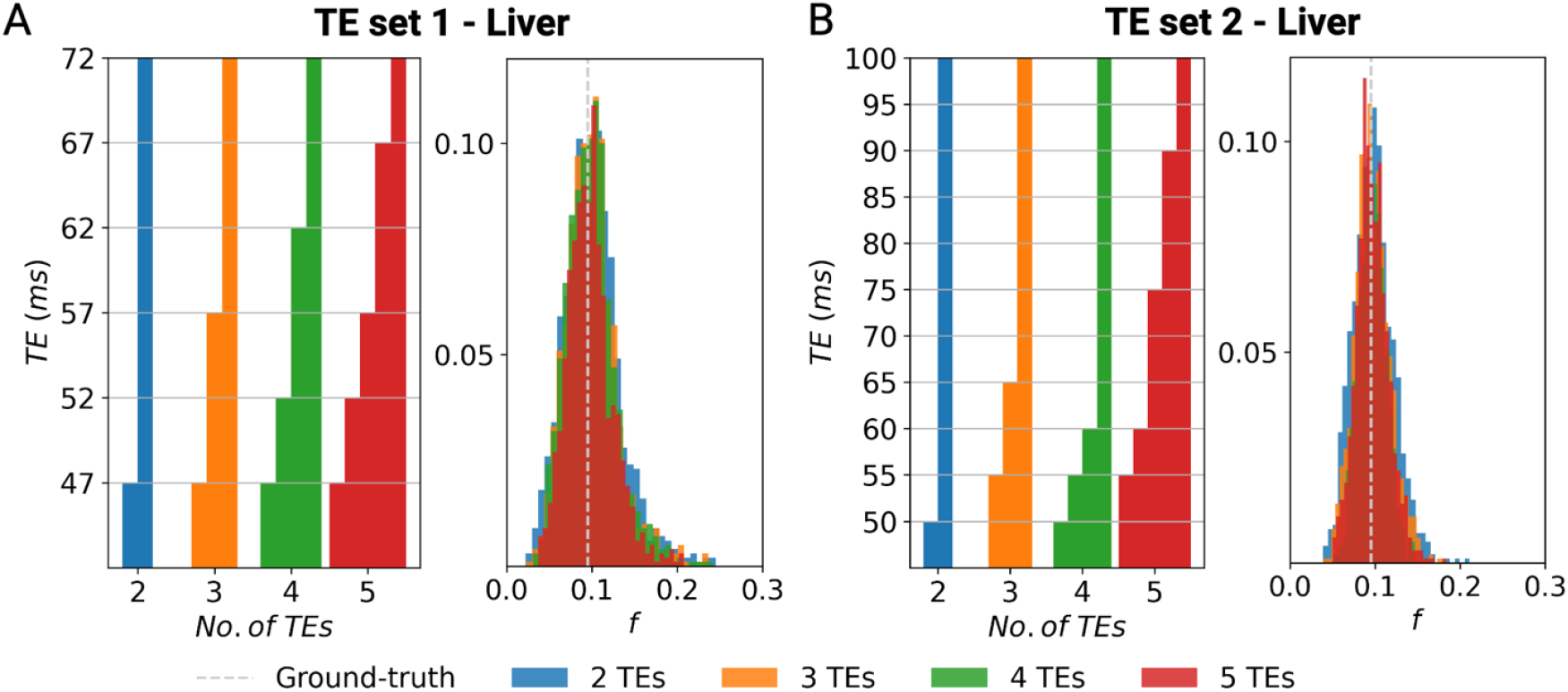
Optimal combinations of TEs (left panel) and the corresponding distributions of pseudo-diffusion volume fraction *f* (right panel) from the 2D T2-IVIM fit of the liver *in silico* data (N_rep_ = 2500, SNR_S0_ = 40) using these optimal TE values in the TE range from 47 ms to 72 ms (TE set 1, A) and from 50 ms to 100 ms (TE set 2, B).

As demonstrated in **Figure 7**, these findings were validated using a subset of the *in vivo* data acquired at the optimal b-values and different TE values from the TE set 1. **Figure 7A** shows the representative *f* maps and the difference maps obtained in the liver, which look very similar for the optimized TE sets. This observation is further confirmed when comparing the distributions of all pixels in the difference maps, as shown in **Figure 8**. Consistent with our *in silico* results, the 8 b-values + three-TE protocol appears to offer a similar accuracy to the more extensive 16-b-values + 6 TE protocol, but with a shorter scan time for a single subject (Figure 8A) and across all subjects (Figure 8B).

**Figure 7.**
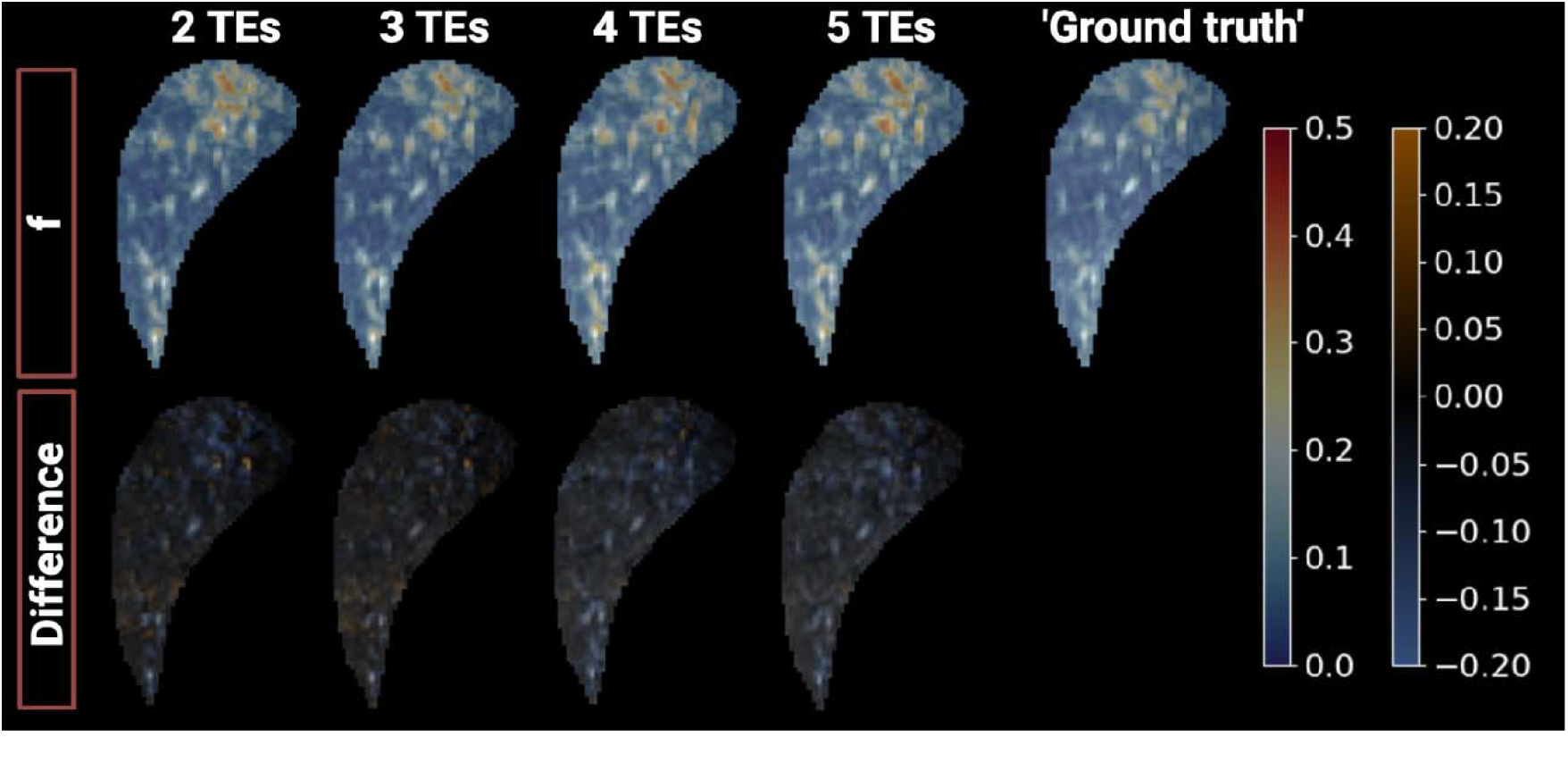
Representative pseudo-diffusion volume fraction *f* and difference maps obtained in the liver of a healthy volunteer from fitting the data to the T2-IVIM model using the optimal TE combinations from the TE set 1 and six b-values. The ‘ground truth’ *f* map was obtained from the 2D fit using all 6 TEs and 16 b-values and was used to calculate the difference maps.

**Figure 8.**
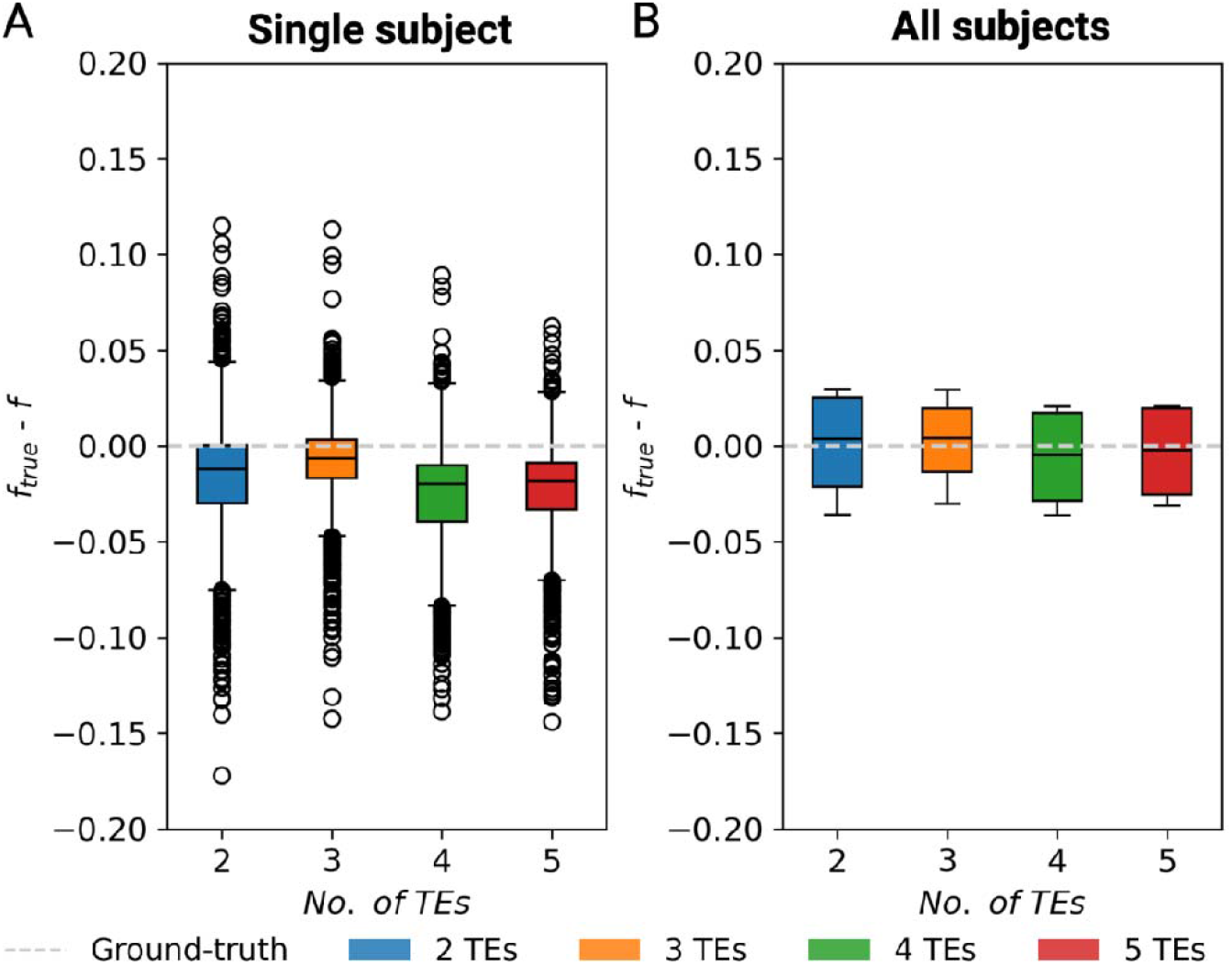
Pixel distributions of the difference between the *f* values obtained with a full *in vivo* protocol (‘ground truth’, *f*_*true*_) and optimized 2, 3, 4, and 5 TEs protocols in a representative subject from Figure 7(A), and the distributions of mean differences in all subjects (B).

## Discussion

In this study, we quantitatively assessed bias in pseudo-diffusion volume fraction when using the traditional IVIM model without accounting for the differences in compartmental T1 and T2 relaxation times. While the T1-dependence of *f* can be eliminated by choosing long TR, the effect of TE should be minimized by applying simultaneous T2-IVIM modeling (combined with the IDEAL fitting approach) using two or three different TE values, if possible. Our *in vivo* findings revealed that the T2-compensation is particularly important when analyzing the IVIM data collected in the liver.

The evidence from this study supports our hypothesis that the large variability^33,34^ in the reported pseudo-diffusion volume fractions across different IVIM studies can partially be explained by the variation in TE used for the DW data acquisition at different centers. This variation in TE results from the common practice, both in clinical and research DWI protocols, to use the minimum TE, which is determined by the gradient hardware, maximum b-value and the diffusion encoding scheme. Furthermore, our results also suggest that age-, physiology- or pathology-related changes in IVIM-derived *f* cannot straightforwardly be interpreted as changes in blood perfusion or other flow-related processes unless no changes in compartmental T2 can be assumed. For instance, higher *f* values measured in older subjects maybe be associated with the age-related increase in liver iron content and thus shorter T2 (contributed by both tissue and blood T2)^35^. In other studies, the extent of reduction in *f* due to perfusion reduction in liver fibrosis could have been overestimated as T2 has also been shown to increase in this pathology^36,37^. Similarly, as T2 and T2* increase with higher local blood oxygenation^38^, *f* is expected to be influenced by the oxygenation status. The use of T2-IVIM modelling can serve as a potential solution in overcoming these issues by providing TE-independent *f* measures to improve the reproducibility in multi-center research settings, and to contribute to better understanding of potentially relevant sources of variation in *f* estimation in various pathologies.

The relaxation time dependence of *f* has been found in several studies conducted in the normal liver^39–41^, normal prostate^42^, pancreas^24^, and locally advanced breast cancer^43^. In general, an increase in *f* with longer TE was observed in all these organs, which is consistent with the results of our numerical experiments and *in vivo* study in the liver. Furthermore, our mean *f* values (between 0.17 ± 0.09 at TE = 47 ms and 0.28 ± 0.11 at TE = 72 ms) measured in the liver are in good agreement with those from previous studies (Table 4 in Führes et al.^40^). Similarly, our mean liver T2_fluid_ of 60 ± 6.8 ms is well within the range of values reported for 1.5 T by Jerome et al.^39^ (T2 = T2 = 77.6 ± 30.2 ms), and notably shorter than the previously reported blood T2 (e.g., 275 ± 50 ms at 3T, with 95% blood oxygenation^23^. As expected, the mean T2 of 26.5 ± 2.2 ms value found in our study is slightly lower but overall comparable with the liver T2 values reported by other groups (e.g., 34 ± 4 ms at 3T^44^, or 42 ± 3 ms at 3T^23^).

To the best of our knowledge, this is the first study to investigate the TE-dependence of the IVIM measures in the kidney. The relaxation-compensated *f* values of 0.17 ± 0.03 and 0.13 ± 0.04 measured in the renal cortex and medulla, respectively, are consistent with literature values^2,25,29,45,46^. The mean T2 values (62.5 ± 14.16 ms and 70.6 ± 21.2 ms in the cortex and medulla, respectively) are close to cortical and medullary T2 values reported by de Bazelaire et al.^44^, and the T2 values (158.7 ± 58.8 ms and 154.0 ± 44.1 ms in the cortex and medulla, respectively) are substantially higher than those of the liver. Interestingly, no clear trend in *f* with increasing TE was found in the TE range between 47 and 72 ms. This can partially be explained by the additional “pseudo-diffusion” effect in the kidney caused by the presence of pre-urine (i.e., glomerular filtrate) flow within the renal tubules (in addition to blood flow in the peritubular capillaries) that introduces a third compartment^47–50^ with distinct relaxation and diffusion properties. In addition, this pre-urine compartment is expected to have a relatively long T2 value, which might not have been sufficiently probed at the relatively short TE values used here. This study complement our prior work in which we showed an increase in *f* with increasing diffusion time at constant TE^25^. A future study with larger sample size, broader range of TE values, and more advanced T2-IVIM modelling (e.g., three-compartment T2-IVIM modeling) is needed to validate these findings.

In outlook on potential clinical applications, optimizing the T2-IVIM imaging protocol to ensure the accuracy of the estimated IVIM parameters while minimizing the acquisition time is of particular relevance. For liver T2-IVIM, Jerome et al.^39^ proposed a clinically feasible 11 min protocol at 1.5 T that consisted of one acquisition with 8 b-values (0 – 800 s/mm^2^) at TE = 62 ms and two additional low b-value scans (0, 10, and 50 s/mm^2^) a two longer TEs (80 and 100 ms). However, this protocol was designed empirically and minimizing the bias in *f* was not the primary goal. Here we propose a numerical framework that optimize the TE values for relaxation-compensated *f* parameter estimation for any given set of b-values, relaxation properties, and expected IVIM parameters. This flexibility enables efficient protocol optimization for various organs and pathologies if rough estimates of the model parameters are known. Our results suggest that a b-TE protocol consisting of two shorter TEs (50 ms, and 55 ms) and one longer TE (100 ms) combined with six b-values recommended by the recent *consensus*^*51*^ might be optimal for liver T2-IVIM if the focus is on estimating the pseudo-diffusion volume fraction. This finding can be explained by the relatively short T2 of the liver (and therefore the need for short TE) and the fact that lower SNR at higher TE values negatively impacts the robustness of the 2D T2-IVIM modelling. Further acquisition time reduction could potentially be achieved by allowing variable numbers and values of b-values for each TE in the optimization process, which will be a subject of our future works. In addition to the protocol optimization, the use of accelerated b-TE acquisition methods based on multi-echo DWI sequences could be considered^52–55^.

To date, only one study has investigated whether correcting for TE dependence in IVIM imaging offers added clinical value^43^. Based on their findings in breast cancer patients, Egnell et al.^43^ concluded that while the corrected model provides more accurate estimates of *f*, the additional scan time needed to acquire the T2-IVIM data may not be justified when considering the clinical benefit. One reason for this could be the overall low reliability of the *f* parameter estimation in their study due to low SNR which required fixing the T2_fluid_ (T2_p_) value for fitting. We addressed this issue in our study by applying the IDEAL fitting approach that was shown to be more robust to poor SNR and to provide more robust IVIM parameter estimates than the conventional fitting methods by exploiting the concept of spatial homogeneity^29,30^. In this work, the T2 and IVIM parameter maps obtained with the IDEAL approach exhibit superior visual quality and reduced noise compared to those produced by the segmented fitting method. Furthermore, as our method does not require fixing any parameters, it may be better suited for studies investigating the added clinical value of the combined information from the compartmental T2 and IVIM maps. However, if the scan time is a limiting factor, our study suggests that diffusion coefficient *D* might be a better potential biomarker than *f* as it does not depend on TE and its coefficient of variation is typically lower than that of D*^27^.

Limitations of this study include the small study population, and the limited range of TE values used in the *in vivo* experiments. Furthermore, the lack of “ground truth” and test-retest data does not allow us to specifically evaluate the accuracy and precision of the T2 and IVIM parameter estimation with our method. This will be pursued in prospective studies in which the proposed technique and the optimized protocol will be applied in a range of hepatic pathologies.

## Conclusion

In conclusion, the difference in T2 between the tissue and fluid compartments introduces a TE-dependent bias in pseudo-diffusion volume fraction when using the traditional IVIM model. This study shows that two-dimensional IDEAL fitting of the extended T2-IVIM model allows removal of this confounding effect and enables simultaneous estimation of the IVIM parameters and compartmental T2 values in the liver and kidney.

## Data Availability

All code and in silico data produced will be available online at https://github.com/stabinska/T2-IVIM.

## Acknowledgements

Financial support by the National Institutes of Health (K99 DK138294, K99 AG080084) and the Deutsche Forschungsgemeinschaft (408765040, 497764939) is gratefully acknowledged.

