## Supplementary Material for "Toward optimized intravoxel incoherent motion (IVIM) and compartmental T2 mapping in abdominal organs"

**Supplementary Material**
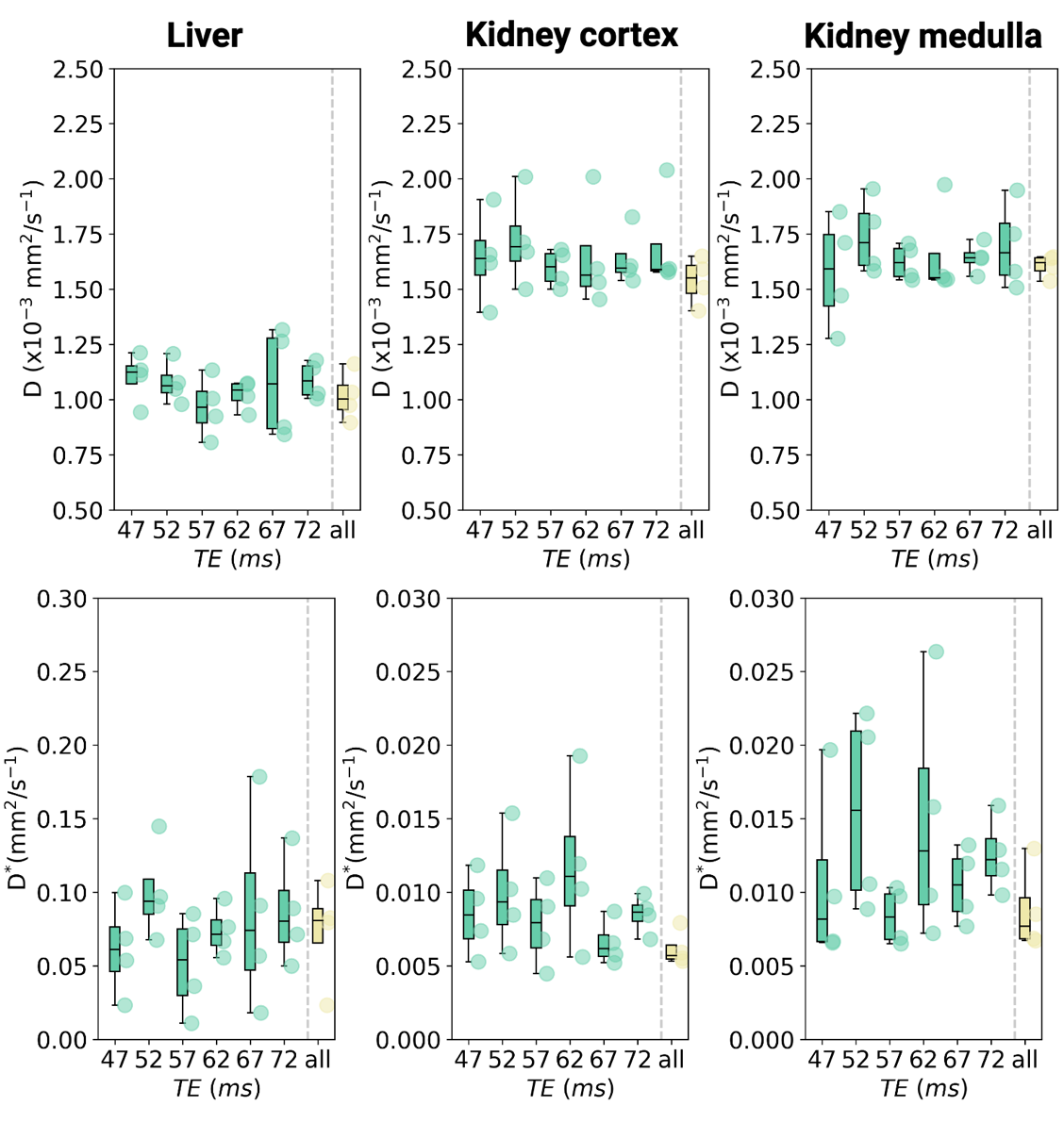

**Figure S1**: Diffusion (D) and pseudo-diffusion coefficients (D*) obtained in all volunteers (N = 4) in the liver, kidney cortex, and kidney medulla using the IVIM modeling for each TE separately (green) and simultaneous 2D T2-IVIM modeling (yellow). The results for each volunteer are displayed as a circle. (*) P < 0.0083.
